# Online Database of Clinical Algorithms with Race and Ethnicity

**DOI:** 10.1101/2023.07.04.23292231

**Authors:** Shyam Visweswaran, Eugene M. Sadhu, Michele M. Morris, Anushka R. Vis, Malarkodi Jebathilagam Samayamuthu

## Abstract

Some clinical algorithms incorporate an individual’s race, ethnicity, or both as an input variable or predictor in determining diagnoses, prognoses, treatment plans, or risk assessments. Inappropriate use of race and ethnicity in clinical algorithms at the point of care may exacerbate health disparities and promote harmful practices of race-based medicine. We identified 42 risk calculators that use race as a predictor, five laboratory test results with different reference ranges recommended for different races, one therapy recommendation based on race, 15 medications with guidelines for initiation and monitoring based on race, and four medical devices with differential racial performance. Information on these clinical algorithms are freely available at http://www.clinical-algorithms-with-race-and-ethnicity.org. This resource aims to raise awareness about the use of race in clinical algorithms and to track the progress made toward eliminating its inappropriate use. The database will be actively updated to include clinical algorithms based on race that were missed, along with additional characteristics of these algorithms.

## INTRODUCTION

Clinical algorithms are tools that aid decision-making in a variety of medical conditions and procedures. Examples include diagnostic calculators that assess the current presence of a disease or condition, prognostic algorithms that predict the future risk of a disease or health outcome [1], treatment guidelines for managing chronic conditions, interpretation recommendations for laboratory test results, and precision directions for medication use [2, 3]. Such algorithms enable standardization of care, increased efficiency, and improved clinical decision-making quality [2].

Some clinical algorithms incorporate the individual’s race, ethnicity, or both as an input variable or predictor. In medicine, race and ethnicity are used to describe certain population characteristics that may have implications for health and health care. While these terms are often used interchangeably, they each have distinct meanings and refer to separate aspects of human identity and ancestry [4]. Race is a categorization system that classifies individuals based on visible physical traits such as skin color, facial features, hair texture, and eye shape. Influenced by historical and political factors, race has been used to classify people into broad groups, such as white, Black, or Asian, among several others. In contrast, ethnicity is a categorization system that groups people based on language, religion, traditions, and other aspects of shared cultural heritage passed down through generations. Shaped by ancestry and geographical location, ethnicity has been used to classify people into culturally distinct groups, such as Hispanic, Chinese, or Navajo, among many others. Despite the distinction between race and ethnicity, we will frequently use the term race to refer to both race and ethnicity for the sake of brevity.

Race is widely used in medicine in studying genetic variations, disease prevalence, treatment responses, and disparities in health and health care. However, using race as a proxy for genetic differences or more broadly biological differences can oversimplify complex health issues and contribute to disparities. Frequent causes of health disparities include, among others, environmental, socioeconomic, health care access, discrimination, and cultural factors. Although clinical algorithms incorporating race are intended to improve health care, they can inadvertently exacerbate racial health disparities in several ways. They may embed racial prejudices and stereotypes arising from historical and societal biases, and the inclusion of race as a predictor can inadvertently reinforce these biases. Medical students are taught to associate race with diseases such as sickle cell anemia, sarcoidosis, or cystic fibrosis, which reinforces the use of race as a proxy for the biological basis of disease [5]. Using race as an alternative for genetic variation oversimplifies the complex interactions between genes, environment, and disease [6]; furthermore, there is more genetic variation within racial groups than across them [7]. Thus, the use of race in clinical algorithms may incorrectly presume genetic and biological differences between racial groups, and may result in incorrect diagnoses or inadequate treatment in minority populations [8]. Further, the use of race in clinical algorithms raises ethical concerns regarding discrimination, equity, and equality of opportunity [9]. It may violate the principles of justice and equality if individuals are treated differently based on their race instead of their individual health requirements.

Race is now widely accepted as a historical and social construct rather than a biological one. As a result of growing recognition that diagnosis and treatment based on race reflect flawed biological and genetic assumptions, the use of race in clinical decision-making has come under increasing scrutiny. Several recent high-profile articles have identified and deemed problematic clinical algorithms that include race in a range of clinical specialties, such as nephrology [10], urology [11], obstetrics [12], and cardiology [5, 13, 14]. Further, medical organizations such as the American Society of Nephrology and the American College of Obstetricians and Gynecologists have recommended the elimination of race from algorithms in current use.

However, it is not clear that the inclusion of race as a predictor to inform clinical decision-making will automatically perpetuate long-standing disparities in health and health care [15]. Till recently, it was assumed that clinical prediction algorithms should include all observed patient variables, biological or not, with predictive power to produce more accurate predictions. A recent study showed that dropping race from algorithms may, in fact, propagate racial discrimination and health care inequities [9]. Thus, the process of eliminating race is more nuanced than removing it indiscriminately from all clinical algorithms.

Our goal in this study was to create an up-to-date database of clinical algorithms that include race as a predictor, so that it will a resource for raising awareness of the use of race in clinical decision making and tracking the progress made toward eliminating its inappropriate use. We conducted a systematic search and analysis of dedicated online resources and the published literature for clinical algorithms and identified and classified those that use race.

## METHODS

Clinical algorithms include risk calculators in diagnostic and prognostic settings, flowcharts, lookup tables, nomograms, and guidelines. A risk calculator uses a mathematical equation or a statistical model to assess the presence of a disease (diagnostic calculator) or predict the risk of future onset of a disease (prognostic calculator), such as assessing current osteoporosis status and predicting future fracture risk associated with osteoporosis. A flowchart is a branching decision tree, such as a diagnostic flowchart for determining the etiology of chest pain. A lookup table enables quick reference of data, such as a table containing energy and nutritional content of various foodstuffs. A nomogram is a graphical tool used for a specific calculation, such as a nomogram of height and weight measurements used to find the surface area of a person. This study focused on identifying and cataloging clinical algorithms such as risk calculators that use race as a predictor, laboratory test results with different reference ranges recommended for different races, therapy recommendations based on race, and medications with guidelines for initiation and monitoring based on race.

### Data sources and search strategy

We identified online resources with clinical calculators using Google search (query: (medical OR clinical) AND calculator). Websites were excluded if there was no contact information, no references (i.e., PubMed), contained only hyperlinks to external websites, described smartphone apps for calculators, or did not implement the calculator for online data entry and result output. From the online resources that met inclusion criteria, we created a list of risk calculators that included race as a predictor with race and had at least one PubMed reference.

We also identified peer-reviewed articles with clinical calculators with race using PubMed search (query: (medical OR clinical) AND calculator AND race AND bias) and retained those articles that contained a table of clinical calculators with race. From the articles that met inclusion criteria, we generated a list of clinical calculators that included race as a predictor. After merging the list of calculators obtained from online resources with the list obtained from peer-reviewed articles, we removed duplicates to create a final list.

We identified medications with guidelines based on race using Micromedex, which is one of the largest web-based pharmacological knowledge bases that provide detailed information on drugs and their clinical significance. We generated independent lists for each of the following keywords: ancestry, descent, ethnicity, heritage, and peoples. After merging the lists and removing duplicates, we further removed medications with no U.S. FDA Drug Label Information, medications with no race-based guidelines in the Drug Label Information, and combination medications if a member of the combination was already included.

We also identified peer-reviewed articles with medical devices with differential racial performance using PubMed search (query: (medical OR clinical) AND device AND race AND bias) and retained those articles that described a medical device with differential performance across races.

### Data extraction

We reviewed the online source and the PubMed reference for each included calculator and extracted pertinent information. The information included the name of the calculator, its purpose, a description of its use of race and rationale for the use of race, the predictors, a PubMed reference, and a description of modifications made to eliminate race after its introduction. In addition, we categorized each calculator based on its intended use and identified its clinical specialty.

We reviewed the FDA Drug Product Labeling for each included medication and extracted pertinent information. The information included the name of the drug, a description of the drug, a description of the use of race and rationale for the use of race, a reference to the FDA Drug Label Information, and the section(s) in Drug Label Information that contained guidelines based on race. In addition, we categorized each medication according to the racial context of its use and identified its clinical specialty.

We reviewed the articles retrieved from PubMed reference for each included device and extracted pertinent information. The information included the name of the device, its purpose, a description of its use, and a description of racial differences in the performance of the device, and a PubMed reference.

### Online database

After identification, data extraction, and validation, we created an online database that provides free access to the results. We plan to regularly update this open-access database as new calculators, medications and other clinical uses of race are identified. We added a submission feature to the database, allowing the community to submit new calculators and medications that our search missed. Before being added to the database, all community submissions will be verified and cross-checked.

## RESULTS

We report separately clinical calculators with race (including risk calculators, laboratory tests, and therapy recommendations), medications with guidelines based on race, and devices with differential racial performance.

### Clinical calculators

We identified 191 online resources with clinical calculators, of which 37 met the inclusion criteria and 208 articles, of which three met the inclusion criteria (Figure 1). After merging the lists of calculators identified from the online resources and the three articles respectively, the final list contained 45 calculators. Detailed information for each calculator is provided in the online database. The public has contributed three new calculators, bringing the total to 48. Detailed information for each calculator is provided in the online database.

**Figure 1.**
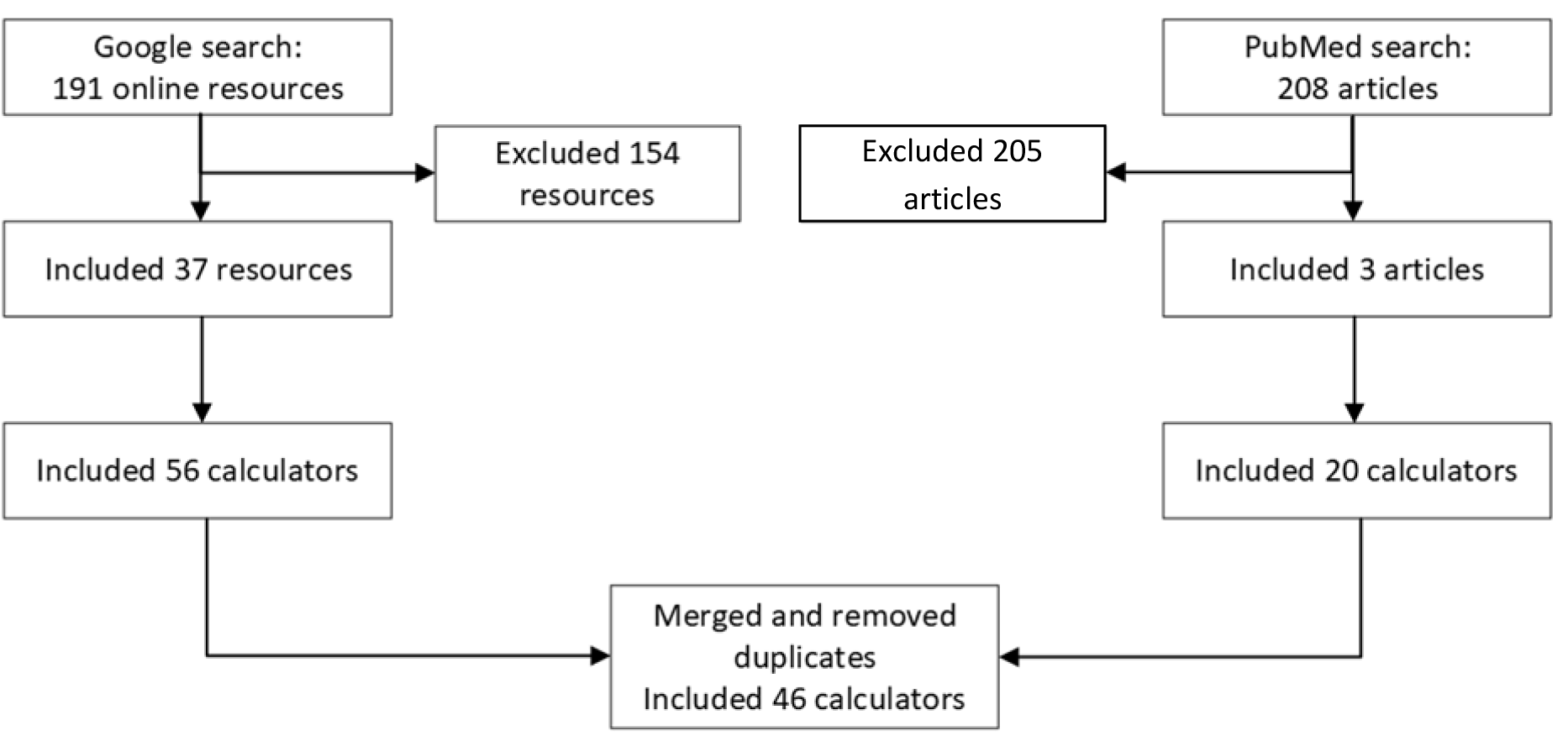
Selection of risk calculators, laboratory tests, and therapy recommendations with race.

Among the online resources, the most comprehensive are MDCalc, UpToDate Medical Calculators, and MDApp. MDCalc is widely used globally, and in the U.S., over 65 percent of physicians use it every month [16]. UpToDate is a widely used database of point-of-care information that also contains a comprehensive calculator resource [17]. MDApp is a U.K.-based company that implements clinical calculators. The MD Anderson Cancer Center has created several cancer-related calculators to predict survival, clinical outcomes, and response to treatments.

Based on intended use, the clinical calculators were categorized into risk calculators, laboratory tests, and therapy recommendations (see Table 1). The calculators were also grouped into ten specialties, including cardiac surgery (1), cardiology (5), endocrinology (3), infectious diseases (4), nephrology (3), obstetrics (13), oncology (10), pulmonology (3), surgery (2), and urology (2). The rationale for the use of race was mostly based on epidemiological data that recorded race, and a statistical analysis of the data found a difference based on race. Seven of the 48 calculators have been modified to exclude race as a predictor (Anemia in pregnancy, ASCVD Risk Calculator, MDRD GFR Equation, Kidney Donor Risk Index (KDRI), Spirometry Reference Value Calculator, Vaginal Birth After Cesarean (VBAC) and UTICalc).

**Table 1.**
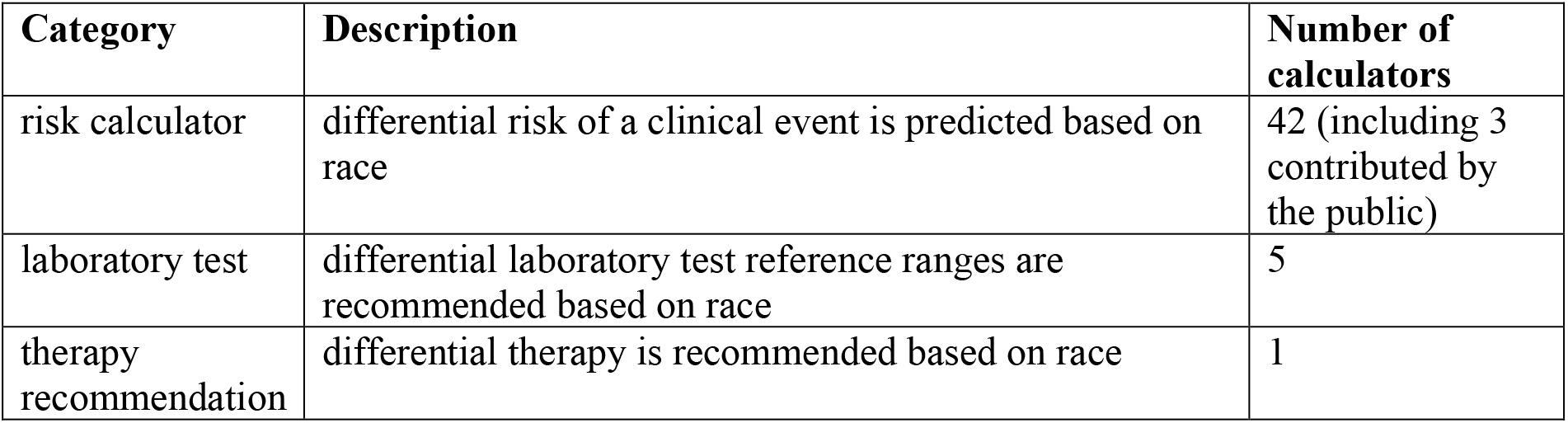
Summary of risk calculators, laboratory tests, and therapy recommendations with race.

| Category | Description | Number of calculators |
| --- | --- | --- |
| risk calculator | differential risk of a clinical event is predicted based on race | 42 (including 3 contributed by the public) |
| laboratory test | differential laboratory test reference ranges are recommended based on race | 5 |
| therapy recommendation | differential therapy is recommended based on race | 1 |

Race and ethnicity are treated as separate predictors in only three calculators (Predict COVID-19 Test Result, Predict Hospitalization Risk for COVID-19 Positive, and Kidney Donor Risk Index); in the majority of the calculators, race and ethnicity are treated as a single variable or only race is included as a predictor. A total of 49 distinct race/ethnicity categories were identified in the calculators. Common categories included white, Black, other, Asian, Caucasian, East Asian, mixed, and South Asian.

### Medications

From Micromedex, we identified 47 (keyword: ancestry), 108 (keyword: descent), 133 (keyword: ethnicity), 17 (keyword: heritage), and 5 (keyword: peoples) medications with potential race-based guidelines. After removing duplicates and applying the exclusion criteria, the final list contained 15 medications for which we verified that the FDA Drug Product Labeling included guidelines based on race (see Figure 2). Detailed information for each medication is provided in the online database. Based on how race was used, medications were categorized into those with race-based indication, race-based dose adjustment, race-based monitoring, and race-based pharmacogenetic screening (see Table 2). The rationale for the use of race mostly came from studies that recorded race and found a statistical difference in genetics or pharmacokinetics based on race.

**Table 2.** Summary of medications with guidelines based on race.

| Category | Description | Number of medications |
| --- | --- | --- |
| race-based indication | medication is recommended for a specific race or races | 2 (isosorbide dinitrate and hydralazine, isoniazid) |
| race-based dose adjustment | differential dosing is recommended based on race | 6 (rosuvastatin, eltrombopag, tacrolimus, warfarin, omeprazole, simeprevir) |
| race-based monitoring | differential frequency of monitoring is recommended based on race | 2 (simvastatin, alvimopan) |
| race-based pharmacogenetic screening | pharmacogenetic screening is recommended for a specific race or races | 5 (pegloticase, rasburicase, carbamazepine, oxcarbazepine, allopurinol) |

**Figure 2.**
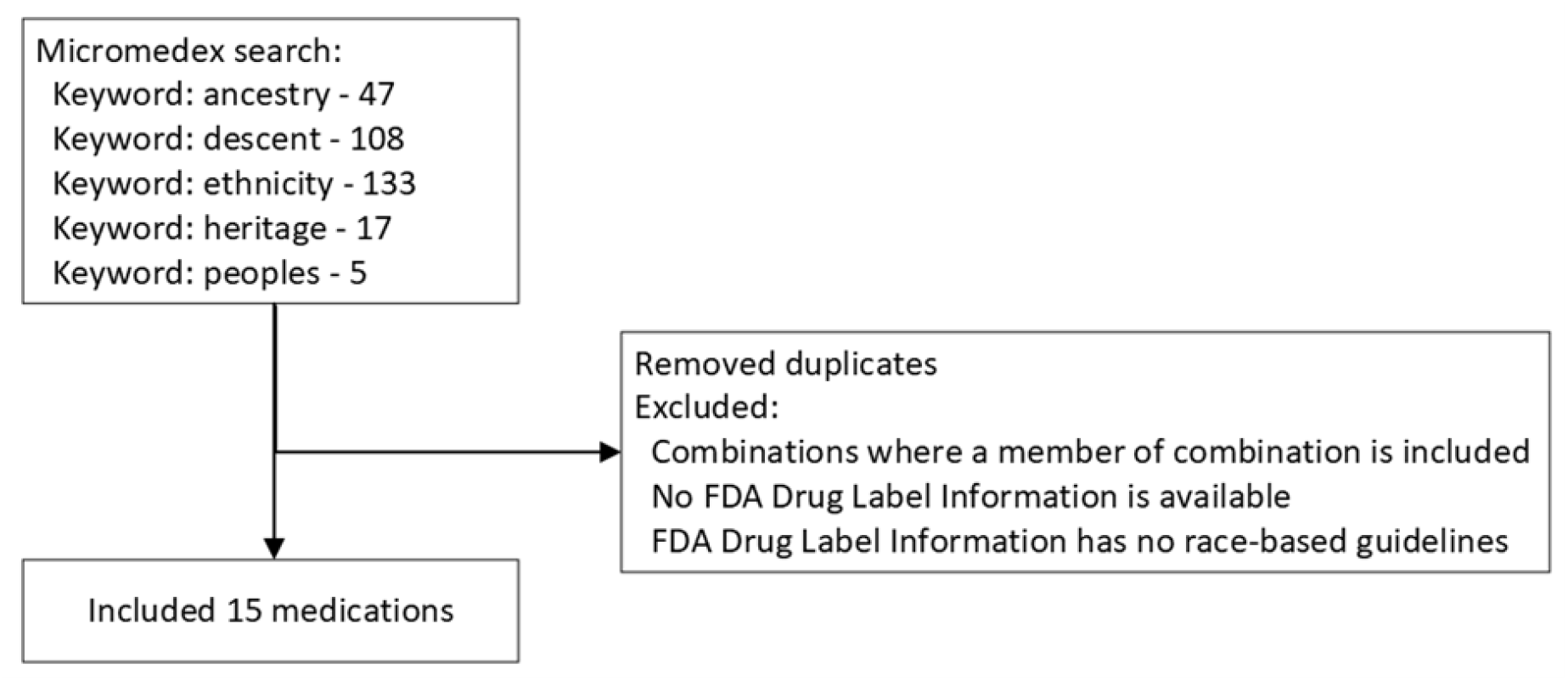
Selection of medications with guidelines based on race.

### Medical devices

We identified 83 articles, of which four met the inclusion criteria (Figure 3). Detailed information for each medical device is provided in the online database and in the Supplementary Information (see Spreadsheet 3). The list of devices with differential racial performance is shown in Table 3. The causes for differential performance arise from differences in skin pigmentation for three devices and hair type for one device.

**Table 3.** Summary of medical devices with differential racial performance.

| Device | Differential racial performance |
| --- | --- |
| Pulse oximeter | Research indicates that pulse oximeters overestimate blood oxygen levels in people with darker skin. |
| Temporal artery (forehead) thermometer | Temporal temperature measurement was associated with a lower likelihood of detecting fever in Black patients than oral temperature. |
| Electroencephalogram (EEG) scalp electrode | For thick curly hair and hairstyles commonly worn by Black people, attaching EEG electrodes optimally can be challenging. |
| Photoplethysmographic sensor | Photoplethysmographic sensors in wearable devices used to detect atrial fibrillation have been linked to poor performance on darker skin tones. |

**Figure 3.**
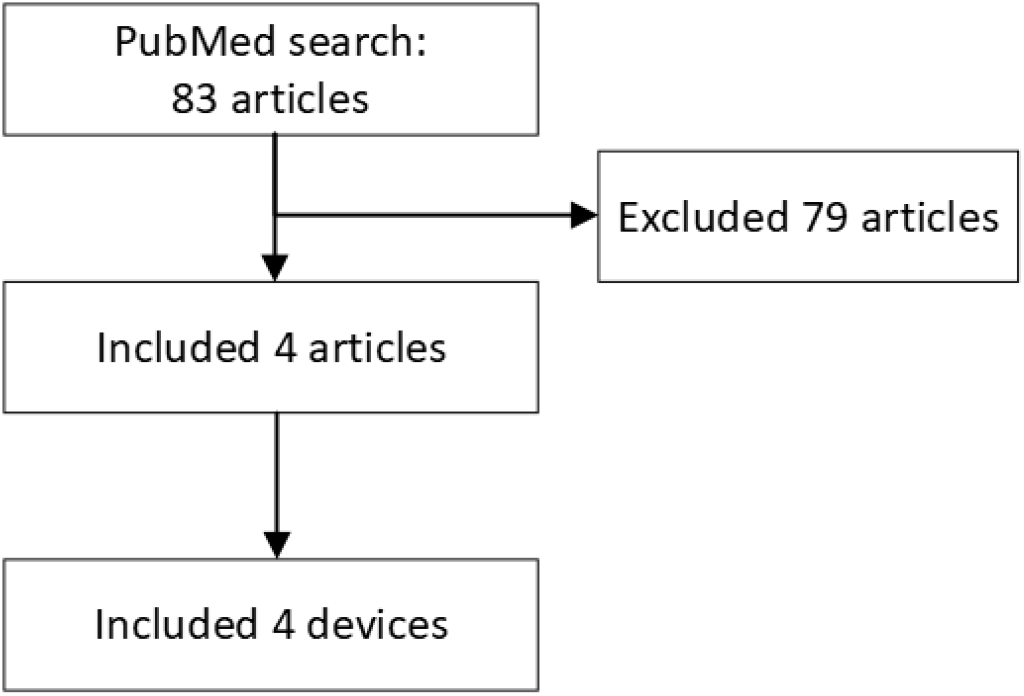
Selection of medical devices with differential racial performance.

## DISCUSSION

Clinical algorithms are embedded in electronic health records, guidelines, and decision support tools, and it is feared that the use of algorithms that include race as a predictor may lead to disparities in health care, especially in racial minority populations. As our understanding of race in medicine evolves, efforts are being made to investigate the role of race in clinical tools. Recently, the Agency for Healthcare Research and Quality conducted a stakeholder review of 18 algorithms based on race in health care [13]. With the beginning of this new era, it is crucial for the medical community to have an up-to-date catalog of clinical algorithms that use race in order to anticipate and measure progress in the antiracist reformulation of these tools. We conducted an exhaustive search of online resources, the scientific literature, and the FDA Drug Label Information and identified 39 risk calculators that use race as a predictor, six laboratory test results with different reference ranges recommended for different races, one therapy recommendations based on race, and 15 medications with guidelines for initiation and monitoring based on race. Information about these 61 entities is available in an online database at http://www.clinical-algorithms-with-race-and-ethnicity.org/.

A key issue is the lack of standardization in the racial categorization systems currently in use. Typically, an optimal categorization system has consistent definitions for universally applicable categories, and the categories are mutually exclusive. A good system is also complete and capable of absorbing entities that have not yet been identified without requiring system revisions. All of these features are lacking in racial categorization systems. There is no single global racial categorization system in use. The Office of Management and Budget in the U.S. defines five racial categories: white, Black or African American, American Indian or Alaskan Native, Asian, and Native Hawaiian or Other Pacific Islander, as well as two ethnic categories: Hispanic and not Hispanic. In the U.K., the Office of National Statistics defines five high-level ethnic groups: “Asian, Asian British, Asian Welsh,” “Black, Black British, Black Welsh, Caribbean or African,” “Mixed or Multiple,” “White,” and “Other ethnic group.” These systems make racial categories difficult to define in practice, and there are no well-defined rules governing what constitutes a race or which race a person belongs to. For example, Caucasians are often called whites or Europeans, even though many Caucasians are neither. Blacks are often called Africans, even though many Blacks are not African. Because racial categories are not mutually exclusive, individuals can belong to multiple races at the same time. Finally, the addition of new racial categories leads to rearrangements of the system. For example, in 1977, the U.S. Census Bureau established four racial categories, including white, Black, American Indian or Alaskan Native, and Asian or Pacific Islander. Two decades later, the Bureau split the Asian or Pacific Islander category into two, namely Asian and Native Hawaiian and Pacific Islander categories. Further, racial categories are used inconsistently across different research papers and data sets. A recent review showed that the U.S. racial categories of white, Black, and Asian mapped to 66, 62, and 49 different racial or ethnic categories, respectively [18]. This lack of standardization leads to ambiguities in operationalizing race-based algorithms for clinical use. For example, guidelines are silent on how race adjustment should be applied to a patient with a white mother and a Black father [14].

Five medications have race-based pharmacogenetic screening recommendations (see Table 2), including testing for glucose-6-phosphate dehydrogenase (G6PD) deficiency in pegloticase and rasburicase, testing for the HLA-B*1502 variant in carbamazepine and oxcarbazepine, and testing for the HLA-B*5801 variant in allopurinol. Race-based pharmacogenetic screening focuses on determining who should undergo specific pharmacogenetic testing so that testing is pursued only for those who are most likely to require it. However, the use of broad racial categories complicates the application of these guidelines in practice. For instance, while the HLA-B*1502 variant is present in over 10% of individuals from Indonesia, Hong Kong, and Vietnam, it is present in less than 1.5% of individuals from Japan and Korea. Therefore, the broad racial category of Asians is inadequate for identifying which patients are at the greatest risk and would most likely benefit from genetic testing [19]. Hence, pharmacogenetic screening recommendations based on race may result in considerable disparities in health care with the potential for adverse clinical outcomes.

Concerns regarding the inappropriate use of race in clinical algorithms have prompted calls for the elimination of race adjustments in clinical algorithms both in the academic literature and by organizations such as the Coalition to End Racism in Clinical Algorithms [20] and the Kaiser Family Foundation [21]. These efforts have resulted in the recent removal of race from four calculators. In 2021, the American College of Obstetricians and Gynecologists eliminated cutoffs for hematocrit levels based on race for screening for iron deficiency anemia in pregnancy [22], data from the Cesarean Registry of the Maternal-Fetal Medicine Units Network was reanalyzed to develop a new Vaginal Birth After Cesarean (VBAC) calculator without race and ethnicity [23], and a Task Force established by the National Kidney Foundation and the American Society of Nephrology recommended the use of an updated eGFR equation without race [24]. In 2022, the original UTICalc calculator was replaced by a race-free calculator with comparable predictive performance [25]. In 2024, the American Heart Association adopted the Predicting Risk of cardiovascular disease EVENTs (PREVENT) [26] as a race-free version of the Atherosclerotic Cardiovascular Disease (ASCVD) Risk Calculator [27], and the American Society of Transplantation adopted an updated race- and ethnicity-free Kidney Donor Risk Index (KDRI) [28].

However, it is critical to thoroughly investigate the impact of race on clinical decision-making and health disparities. While replacing race in clinical decision-making with a person’s unique genetic makeup, environmental factors, and other relevant factors is obviously preferable, including race in understanding health and healthcare disparities may still be prudent [29]. Race is often correlated with health disparities due to various factors such as socioeconomic status, access to healthcare, environmental conditions, and historical systemic racism. Removing race from algorithms without addressing these underlying disparities may overlook important risk factors and perpetuate health inequities. And completely ignoring race could lead to an incomplete understanding of health outcomes for marginalized communities. Some diseases and genetic variations are known to have higher prevalence rates among specific racial or ethnic groups. For instance, sickle cell anemia is more common in individuals of African or Mediterranean descent, and genetic mutations causing G6PD deficiency are more common in persons of African, Asian, and Mediterranean descent. In such instances, using race for screening may be acceptable in the interim until a better biological substitute is developed. With rapid progress in precision medicine, it is preferable to avoid race and to use suitable blood and genetic tests. And as genetic tests, including multigene panels and whole genome sequencing, become more affordable, race-based pharmacogenetic screening will become obsolete. The predictive models underlying clinical algorithms are frequently derived from datasets that were assembled from processes of clinical care. These datasets may contain racial biases due to historical disparities in healthcare access and diagnosis. Simply removing race without addressing the underlying biases might perpetuate or amplify existing inaccuracies, leading to misdiagnoses and inappropriate treatment decisions.

There are limitations to our approach. We concentrated on a narrow subset of the extensive clinical usage of race. Beyond risk calculators that use race, differential reference ranges based on race for laboratory test results, and guidelines for medications with guidelines based on race, race is often used to distinguish among variations in physiological processes, genetics, behavior, and cultural characteristics [30]. In the future, we intend to add uses of race outside of calculators and medication guidelines. We did not include the original online implementations of calculators that were subsequently modified to exclude race as a predictor because the original implementations were not available. The future inclusion of race-based implementations of these algorithms will be useful for historical reasons as well as to evaluate if the revised formulations are indeed less biased. We did not include information on the potential harm or equity concerns related to the use of race, because it is currently unclear if the inclusion of race indeed propagates disparities in all such algorithms. Some authors have pointed out that under certain frameworks of social utility and fairness, all individuals are served better when clinical decisions are guided by all predictors, including race. Further, the particular use of an algorithm may inform if race should be included or not. For example, a recent simulation study showed that removing race from diagnostic algorithms could make health care inequities worse, while excluding race in prognostic algorithms that help decide how to allocate resources can mitigate inequities.

The database will be actively updated to include additional clinical algorithms with race and clinical uses of race. In the future expansion of the database, we want to offer additional details of algorithms, such as the mathematical equations and statistical models. In addition, we intend to include descriptions of the datasets from which the algorithms were derived and, if possible, provide access to the datasets themselves.

## Data Availability

All data produced are available online at http://www.clinical-algorithms-with-race-and-ethnicity.org/.

http://www.clinical-algorithms.org-with-race-and-ethnicity.org

